# Anxiety, brain structure and socioeconomic status in middle-aged and older adults

**DOI:** 10.1101/2025.04.14.25325776

**Authors:** Sasha Johns, Caroline Lea-Carnall, Nick Shryane, Asri Maharani

## Abstract

Anxiety disorders confer a significant global burden, impacting mental health, quality of life, and well-being, particularly in aging populations. Prior studies showed the association between anxiety and abnormal brain structure, and socioeconomic status (SES) is linked with both anxiety and brain structure. However, limited research considers the interaction between these three factors, particularly in older adults. Multivariate regression analysis was conducted using 27,563 participants from the UK Biobank to investigate the relationship between anxiety and brain volume in 30 cortical and subcortical regions for the whole sample and separately by sex. It then investigated whether SES affects this relationship. Five out of 30 brain regions had significant negative associations with anxiety in the whole population. These were the thalamus, gyrus, insular cortex, supramarginal gyrus and precentral gyrus, but these relationships were abolished in each area apart from the precentral gyrus when SES was included in analyses. For females, no significant associations were found. For males, lower volume in the precentral gyrus was significantly associated with higher anxiety, but this relationship was no longer significant when SES was considered. The precentral gyrus was most robustly associated with anxiety across analyses. However, participants with conditions that may affect the brain were not excluded and time between anxiety assessment and brain scans varied widely across participants. Information about past SES during brain development is also not available. To conclude, the relationship between anxiety and brain volume is affected by SES. Clinicians and researchers should take this into account when working with imaging data.

## Introduction

Anxiety disorders are one of the most prevalent mental health conditions, affecting 301.4 million people globally (GBD 2019 Mental Disorders Collaborators, 2022). Anxiety disorders are characterised by anxiety, fear and associated behavioural abnormalities (Yu et al., 2025) and can significantly impact individuals’ psychosocial functioning and quality of life (Cramer et al., 2005; Mendlowicz & Stein, 2000; Olatunji et al., 2007). The effect of anxiety disorders on quality of life is also independent of sociodemographic factors, somatic health and other mental disorders (Cramer et al., 2005).

While anxiety can affect individuals across the lifespan, and some research suggests that rates may be lower in older age (Byers et al., 2010; Jorm, 2000; Lenze & Wetherell, 2011a; Schaub & Linden, 2000; Wolitzky-Taylor et al., 2010), there are different considerations for the understanding and treatment of this condition in older adults (Bryant et al., 2013; Flint, 2005; Flint et al., 1996; Roberts et al., 2017). As the global population ages, with older adults increasing in both number and proportion in every country (World Health Organization, 2024), maintaining mental and physical health in this group is vital for individual wellbeing, healthcare systems and the economy (Curran et al., 2020). Older adults face distinct challenges, including medical comorbidities, financial strain, and reduced social capital, which negatively impact wellbeing and quality of life (Noto, 2023; Ribeiro et al., 2020; Roberts et al., 2017). Anxiety, which may be more prevalent than depression in this population (Andreas et al., 2017; Bryant et al., 2007; Singleton et al., 2003; Therrien & Hunsley, 2012), has also been shown to significantly contribute to self-reported quality of life (Sarma & Byrne, 2013; Sousa et al., 2017). Despite its prevalence and impact, research on anxiety in older adults and its relationship with brain structure remains limited compared to depression (Blay & Marinho, 2012; Bryant et al., 2007; 2013, Johns et al., 2024; Pachana et al., 2007).

Understanding and treating anxiety in older adults requires consideration of its unique aspects. Older adults may experience age-specific anxiety symptoms such as fear of falling and health anxiety (El-Gabalawy et al., 2013; Gagnon et al., 2005; Howland et al., 1993; 1998; MacKay et al., 2021; Roberts et al., 2017), and anxiety may manifest as somatic complaints in this age group (Fuentes & Cox, 1997). It is also associated with higher rates of medically unexplained symptoms, increased healthcare use, chronic medical conditions, and physical disability (Porensky et al., 2009; Sareen et al., 2006). Barriers to diagnosis in this group include misconceptions that anxiety symptoms are a normal part of ageing, a tendency for older adults to minimise their symptoms, misattribution of anxiety symptoms to physical conditions, issues with remembering symptoms, and difficulty expressing that they are experiencing anxiety, potentially due to stigma (Lenze et al., as cited by Wolitzky-Taylor et al., 2010; Roberts et al., 2017). Changes in the presentation of generalised anxiety disorder (GAD), such as decreased autonomic arousal and clinical anxiety but increased worry and fatigue, further complicate recognition (Nilsson et al., 2019). These challenges often result in missed diagnoses and untreated anxiety despite evidence that this population respond well to evidence-based psychotherapy (Lenze & Wetherell, 2011b).

Anxiety is a complex disorder seemingly caused by a combination of genetic (Ask et al., 2021; Demirkan et al., 2011; Gottschalk & Domschke, 2017; Hettema et al., 2005; Purves et al., 2020; Thorp et al., 2021), environmental (Kendler et al., 2011; Ridley et al., 2020; Sahle et al., 2024), psychological (Narmandakh et al., 2021; Struijs et al., 2021), and biological factors (Kundakovic & Rocks, 2022; Łoś & Waszkiewicz, 2021; Martin et al., 2009; Simkin, 2019). It is associated with structural alterations in brain regions involved in emotion regulation, fear processing, and stress response. The amygdala, which plays a crucial role in the processing of emotional stimuli, specifically fear, shows both increases and decreases in volume associated with anxiety levels (Liu et al., 2024; Schienle et al., 2011; Suor et al., 2020; Warnell et al., 2018;). Similarly, volumetric increases and decreases in the prefrontal cortex, responsible for emotion regulation, executive control and fear processing, are associated with anxiety (Chen et al., 2020; Schienle et al., 2011; Syal et al., 2012; Zhao et al., 2017). Reduced hippocampal volume, also involved in emotion processing, is consistently associated with anxiety throughout adolescence and adulthood (Bremner, 2004; Lipschutz et al., 2024; Logue et al., 2018; Mueller et al., 2013), although one study found increased hippocampal volume in older adults with GAD (Baksh et al., 2021).

Social factors such as socioeconomic status (SES), neighbourhood sociodemographic characteristics, social isolation and childhood trauma are related to anxiety (Chen et al., 2019; Kuzminskaite et al., 2021; Leigh-Hunt et al., 2017; Motoc et al., 2019). SES specifically has been shown to be strongly related to a variety of mental health issues. SES refers to a range of indicators that reflect an individual’s social position or status, most commonly including education, social class, and income (Darin-Mattson et al., 2017). In England, individuals in the lowest 20% income bracket are 2-3 times more likely to develop mental health problems than those in the highest 20% (Marmot, 2010). Individuals with mental health problems are less likely to perform well in school (Schulte-Körne, 2016) and have an increased risk of unemployment (Egan et al., 2016). Measures of SES including unemployment, low income and 12 years or fewer in education were associated with higher scores on the General Anxiety Disorder-7 (GAD-7) questionnaire (Nunes et al., 2022).

Additionally, children in low SES families exhibit increased anxiety, mediated by higher parental anxiety and associated with elevated pre-bedtime cortisol (Zhu et al., 2019). Finally, a link has been established between social factors and brain structure. Infants from low-income families show reduced grey matter volume (GMV) in frontal and parietal lobes and slower trajectories of brain growth during infancy and early childhood (Hanson et al., 2013). Childhood SES is also associated with hippocampal volume in adulthood, with higher SES being linked with increased hippocampal volume (Staff et al., 2012), suggesting a link between early life circumstances and brain development. Exposure to poverty is associated with reduced GMV, white matter volume (WMV) and hippocampal and amygdala volume (Luby et al., 2013). Notably, the relationship between poverty and hippocampal volume was mediated by caregiver support or hostility, and stressful life events (Luby et al., 2013), highlighting potential causal pathways and therefore opportunities for intervention to reduce the impact of poverty.

Despite the recognition of abnormal brain structure in anxiety and the relationship between anxiety and SES, and SES and brain structure, research integrating both biological and social factors in older adults is limited. Johns et al. (2025) examined the relationship between depression and brain structure and found that the inclusion of SES changed this relationship. Since depression and anxiety are highly comorbid yet distinct disorders, and research is lacking in anxiety, it would be interesting to first identify brain regions associated with anxiety in older adults and then assess how SES influences these relationships.

Building on our previous study involving depression (Johns et al., 2025), and our systematic review highlighting an urgent need for more research into anxiety and brain structure in middle-aged and older adults (Johns et al., 2024), this study examines the relationship between anxiety and brain structure in the UK Biobank (UKB). It addresses a critical research gap by exploring this relationship in older adults and is novel in considering the impact of SES on these associations.

## Methods

The UKB is a large-scale biomedical population study including over 500,000 participants with an age range of 40-69, recruited since 2006. In addition to being the largest imaging project in the world, with cross-sectional neuroimaging scans on over 60,000 participants, it has rich sociodemographic, socioeconomic and mental health data. Our study included a subset of 27,563 participants, selected as they had structural imaging, anxiety (GAD-7) score and socioeconomic data, and ongoing consent to participate.

Brain magnetic resonance imaging (MRI) data were collected from three dedicated imaging centres, all equipped with identical 3T Siemens Skyra scanners and 32-channel head coils. The current study employed the T1-weighted images which were acquired at 1mm isotropic resolution using a three-dimensional (3D) magnetisation-prepared rapid-acquisition gradient-echo (MPRAGE) sequence (Miller et al., 2016). The preprocessing of the structural images involved removing the face, extracting the brain, aligning it to the standard MNI15 brain template, and applying non-linear warping to this template (Miller et al., 2016).

Tissue-type segmentation was performed using FAST (FMRBIB’s Automated Segmentation Tool, Zhang et al., 2001) to estimate segmentations for cerebrospinal fluid (CSF), grey matter and white matter (Alfaro-Almagro, 2018). T1-based image-derived phenotypes (IDPs) were calculated for the volumes of major tissue types across the whole brain and specific structures (Miller et al., 2016). The full protocol has been detailed by Miller et al. (2016). This study specifically utilised cortical and subcortical brain volume IDPs (see supplementary materials for UKB IDP Field IDs).

Cortical regions were combined as per Johns et al. (2025), originally based on Harris et al. (2022). Smaller cortical regions were combined to make 23 regions. There was some mismatch between the cortical regions described in Harris et al. (2022) and those in the UKB. These were grouped so they matched Harris’ method as closely as possible, as per Johns et al (2025). Regional groupings are outlined in the supplementary materials.

Hemispheres were combined to make a single region e.g. the right and left inferior frontal gyri (IFG) were combined to make a single total inferior frontal gyrus variable. We chose to combine hemispheres in the absence of a compelling reason to examine them separately. Regions that were split further (e.g. with anterior or posterior divisions), were also combined to make a single region (see supplementary materials).

Anxiety was measured by using the GAD-7 scale from the UKB online follow-up mental health questionnaire (UKB codes 20505, 20506, 20509, 20512, 20515, 20516, 20520). The questions are:

Over the last 2 weeks, how often have you been bothered by any of the following problems?

a. Feeling nervous, anxious or on edge
b. Not being able to stop or control worrying
c. Worrying too much about different things
d. Trouble relaxing
e. Being so restless that it is hard to sit still
f. Becoming easily annoyed or irritable
g. Feeling afraid as if something awful might happen

The seven GAD-7 questions were combined to give an overall GAD score. Questions all used the following scoring:

1. Not at all
2. Several days
3. More than half the days
4. Nearly every day

Responses were scored one point for “not at all” through to four points for “nearly every day” and were added to give an overall GAD-7 score between 7 and 28. Responses “prefer not to answer” (−818) were coded as missing.

SES was comprised of education and income. Education (UKB data field 6138) is defined as the highest qualification the individual holds. We dichotomised individuals as either “college or higher” or “not higher education”, which was the reference category. Income (UKB data field 738) is defined as the average total household income in GBP before tax and is split into five categories: Less than 18,000; 18,000 to 30,999; 31,000 to 51,999; 52,000 to 100,000; and Greater than 100,000.

Ethnicity (UKB data field 21000) was a binary variable where codes for 1001 (white British), 1002 (white Irish) and 1003 (any other white background) were combined to create the “white” category (coded 1), and other ethnicities were combined and coded 0 as “not white.”

Sex in the UKB is defined as biological sex, is split into male and female, and does not account for gender identity.

Multivariate regression analyses were conducted in RStudio (Version 2023.12.1+402) to examine the relationships between both cortical and subcortical volumes and anxiety in six models. Model 0 examined the interaction effects between sex and anxiety for each brain region to determine whether the analyses should be run separately by sex. Model 1 examined the whole population and included age, sex and ethnicity as covariates. Model 2 further adjusted for education and income, also in the whole population. These analyses were then conducted separately for females and males, with Model 3 and 5 being as Model 1 but for females and males respectively and Models 4 and 6 being as Model 2 but for females and males respectively. These six models were run on each of the 30 brain regions (23 cortical and seven subcortical), making 180 models in total. Each model had regional volume as the outcome, with the predictors varying as per the description above. P-values were Bonferroni-corrected for the 30 brain regions.

**Table 1:**
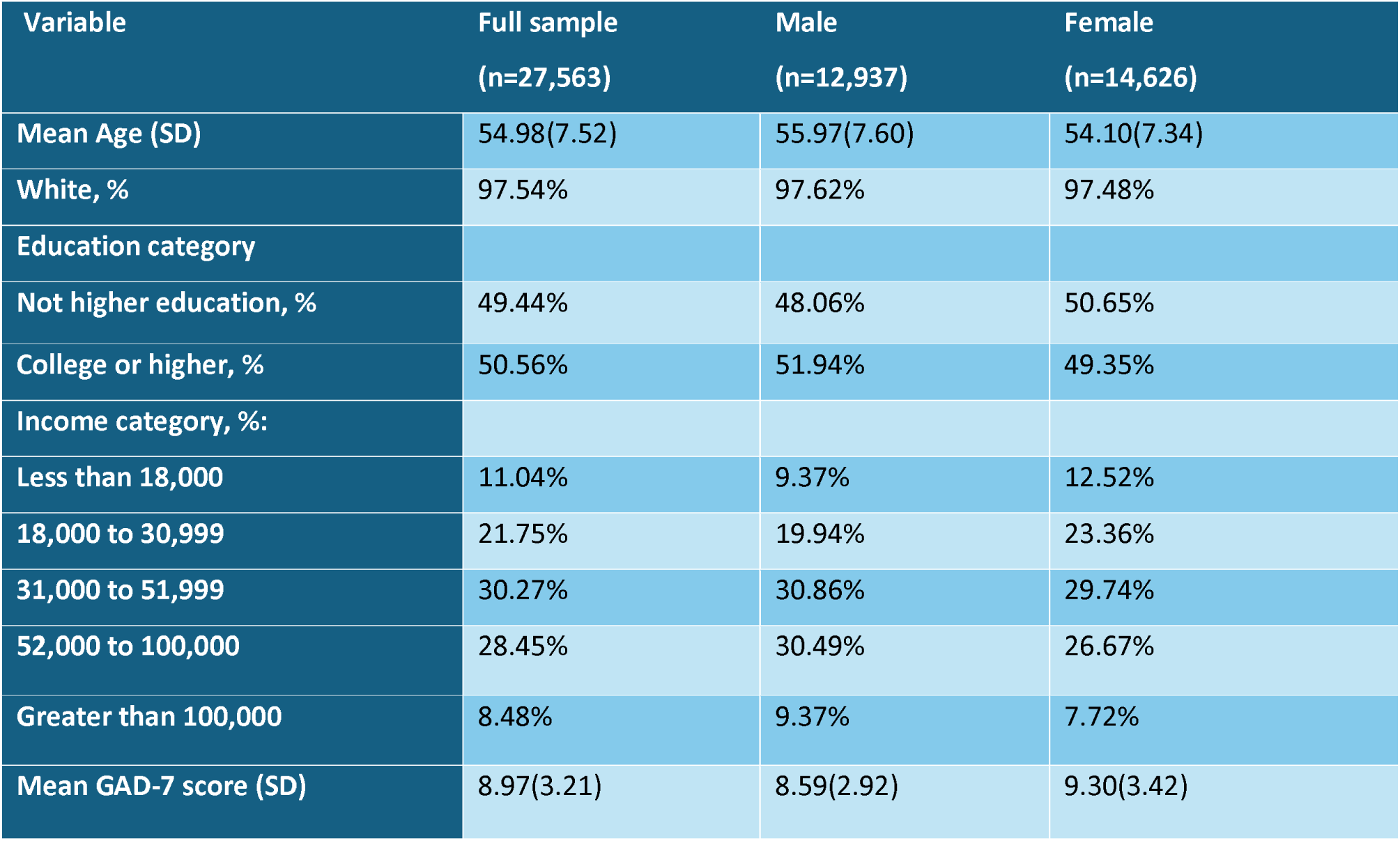
Descriptive statistics for UK Biobank participants included in analyses.

| Variable | Full sample<br>(n=27,563) | Male<br>(n=12,937) | Female<br>(n=14,626) |
| --- | --- | --- | --- |
| Mean Age (SD) | 54.98(7.52) | 55.97(7.60) | 54.10(7.34) |
| White, % | 97.54% | 97.62% | 97.48% |
| Education category |  |  |  |
| Not higher education, % | 49.44% | 48.06% | 50.65% |
| College or higher, % | 50.56% | 51.94% | 49.35% |
| Income category, %: |  |  |  |
| Less than 18,000 | 11.04% | 9.37% | 12.52% |
| 18,000 to 30,999 | 21.75% | 19.94% | 23.36% |
| 31,000 to 51,999 | 30.27% | 30.86% | 29.74% |
| 52,000 to 100,000 | 28.45% | 30.49% | 26.67% |
| Greater than 100,000 | 8.48% | 9.37% | 7.72% |
| Mean GAD-7 score (SD) | 8.97(3.21) | 8.59(2.92) | 9.30(3.42) |

## Results

In Model 0, examining the interaction terms between sex and anxiety for each brain region, the male by anxiety scores terms were significant for increased volume in the superior parietal lobule, the parahippocampal gyrus, the thalamus, the putamen, the pallidum, the hippocampus and the accumbens. These findings suggest significant differences in the relationship between brain volume and anxiety for males and females in these regions, justifying the need to run the analyses separately by sex.

Higher anxiety scores were significantly associated with lower regional volume in five out of 30 (13.33%) brain areas when we adjusted for age and sex in Model 1 (turquoise dots and lines in Fig. 1). However, these relationships only remained significant in one region, the precentral gyrus (b=-9.32, p<.001), when we included education and income in Model 2 (orange dots and lines in Fig. 1.). This suggests that the relationships between anxiety and volume in the three brain areas that were no longer significant (the supramarginal gyrus; the insular cortex; the parahippocampal gyrus; and the thalamus) were potentially confounded by education and income.

**Figure 1:**
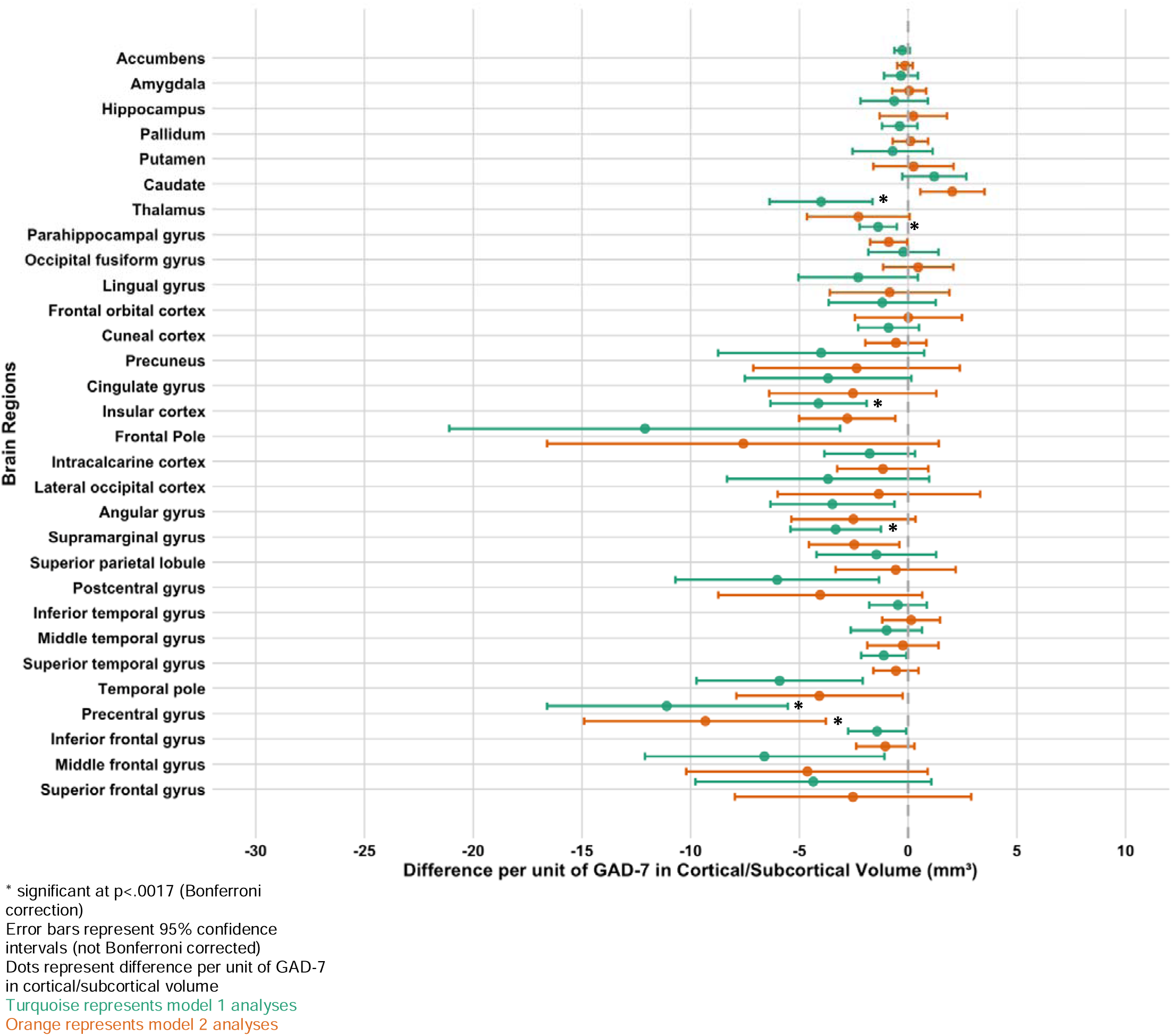
Models comparisons of associations between anxiety and brain regions for the whole population.

Looking at females separately, there were no significant relationships between brain regions and anxiety scores (Model 3, turquoise dots and lines and Model 4, orange dots and lines, Fig. 2).

**Figure 2:**
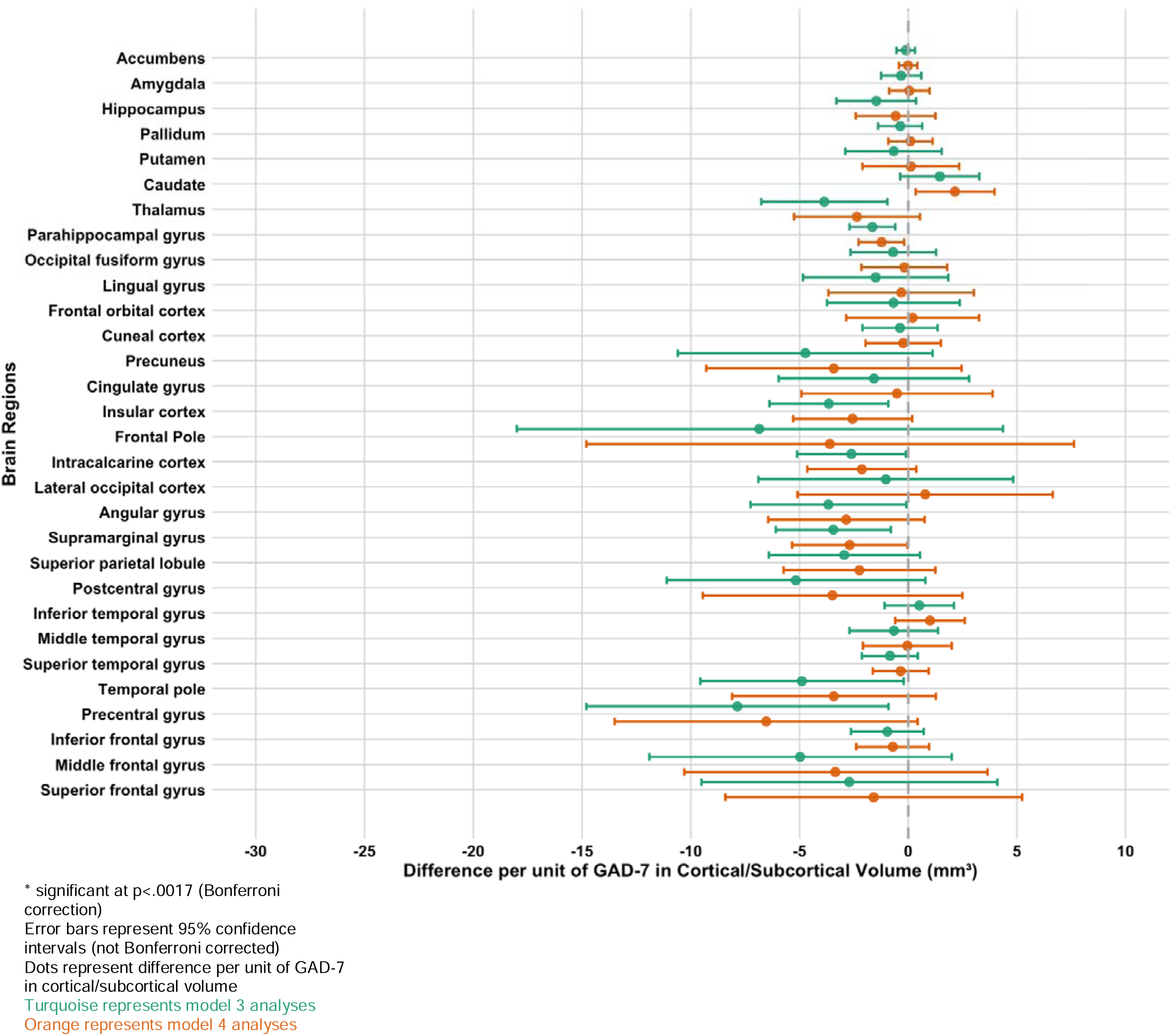
Model comparisons of associations between anxiety and brain areas for females.

Among males, there was only one region that had a significant association with anxiety scores when only age and sex were included in the analysis (Model 5, turquoise dots and lines, Fig. 3). This was the precentral gyrus, and it was negatively associated with anxiety scores in Model 5 (b=-16.10, p<.001). This relationship became non-significant when education and income were included in the model in Model 6 (orange dots and lines, Fig. 3). See supplementary table S1 for results of all models.

**Figure 3:**
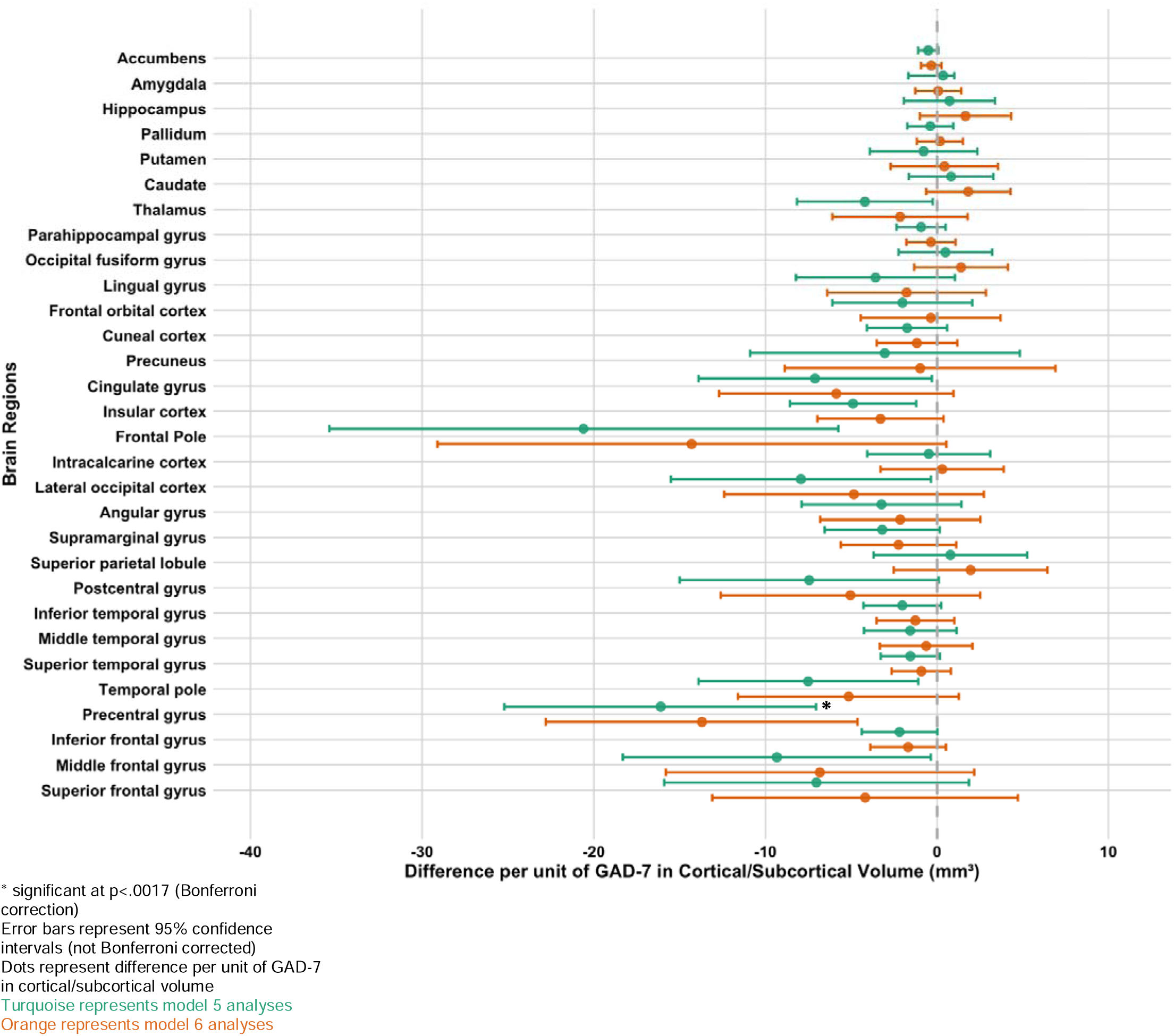
Model comparisons of associations between anxiety and brain areas for males.

## Discussion

A significant negative relationship was found between anxiety and brain volume in multiple brain regions in the UKB dataset. All five brain regions showing significant effects demonstrated negative effects. In almost all brain regions, introducing SES into the model weakens the relationship between anxiety and brain volume, and in four of the five previously significant associations, the effect of anxiety becomes non-significant after including SES.

### Anxiety and Brain Structure

The precentral gyrus exhibited the most consistent association with anxiety scores, showing a negative relationship both before and after controlling for SES in the whole population, and in males when SES was not included. This finding aligns with prior research indicating reduced precentral gyrus volume in individuals with anxiety (Makovac et al., 2016; Shang et al., 2014) and with evidence linking larger volumes in this region to lower risk of having an anxiety disorder in children (Hammoud et al., 2024). Some studies also suggest increased structural integrity of the precentral gyrus in anxiety in terms of thicker cortex and larger GMVs (Gold et al., 2017; Strawn et al., 2013). Functionally, this region is the motor centre of the brain and central to voluntary movement (Banker & Tadi, 2025). It also plays a role in response inhibition by suppressing motor responses as needed (Padmala & Pessoa, 2010; Ray Li et al., 2006; Takeyama et al., 2022; Zhang et al., 2017), with deficits in response inhibition implicated in anxiety (Grillon et al., 2017; Pacheco-Unguetti et al., 2012; Xia et al., 2020; Zhang et al., 2019). It has been suggested that response inhibition may be a potential behavioural marker for clinical anxiety (Grillon et al., 2017). The precentral gyrus is also involved in working memory (Ren et al., 2019; Yue et al., 2019; Emch et al., 2019), and anxiety has been associated with reduced working memory capacity (Lukasic et al., 2019; Moran, 2016; Owens et al., 2012; Vytal et al., 2013) with a bidirectional relationship proposed (Petkus et al., 2017). Additionally, its role in spatial attention (Smith et al., 2010; Yi & Kim, 2020) is relevant, given the contradictory evidence regarding anxiety-related alterations in spatial attention, with some studies suggesting suppressed spatial attention in anxiety (Bachmann et al., 2024; Keller et al., 2022; Kim, 2024; Vytal et al., 2013) and one suggesting improved spatial attention (Caparos & Linnel, 2012).

The supramarginal gyrus displayed a negative association with anxiety scores when SES was not included. This aligns with prior findings of reduced supramarginal gyrus GMV in individuals with GAD (Makovac et al., 2016). This region is implicated in emotion recognition, with the volume of the right supramarginal gyrus being significantly associated with emotion recognition score (Wada et al., 2021) and age-related declines in emotion recognition (Karl & Rohe, 2023). Emotion recognition is also impaired in social anxiety disorder (SAD, Baez et al., 2023), potentially explaining the observed association. The supramarginal gyrus is also involved in theory of mind (Paul et al., 2021), which refers to the recognition that others possess internal mental states—such as intentions, desires, beliefs, perceptions, and emotions—that may differ from one’s own and influence their actions and behaviours (American Psychiatric Association, 2018), another cognitive domain affected by anxiety, with impairments noted in individuals with SAD (Alvi et al., 2020; Baez et al., 2023; Washburn et al., 2016).

The insula also showed a negative association with anxiety when SES was not accounted for, consistent with evidence of reduced insular GMV in individuals with GAD and SAD (Atmaca et al., 2021; Kawaguchi et al., 2016; Moon et al., 2015). The insula is involved in sensory and affective processing as well as higher-level cognition (Uddin et al., 2017). The observed relationship may be explained by anxiety-related disruptions in interoceptive prediction signals, particularly an increased expectation of potentially aversive bodily states. This heightened prediction contributes to increased anxious affect, with the anterior insula playing a central role in this process (Paulus & Stein, 2006). Similarly, the parahippocampal gyrus showed a negative association with anxiety score in the whole population when SES was not included. Given its location near to the hippocampus, a region involved in emotion processing and often related to anxiety within the literature, this is perhaps unsurprising.

However, the current study did not find a significant relationship between hippocampal volume and anxiety scores. The parahippocampal gyrus plays a key role in visuospatial processing and episodic memory (Aminoff et al., 2013; Davachi et al., 2003; Diana et al., 2010; Epstein & Kanwisher, 1998; Mullally & Maguire, 2011; Stevens et al., 2011). Threat-induced anxiety has been linked to disruptions in visuospatial memory accuracy, with differences in physiological measures of anxiety mediating the degree of disruption seen (Shackman et al., 2006), and impairment of consolidation of global visuospatial information (Cody et al., 2014). Alterations in episodic memory are also characteristic of anxiety disorders (Airaksinen et al., 2005; Pajkossy et al., 2017; Zlomuzica et al., 2014) with anxiety symptoms predicting episodic memory decline in cognitively healthy older adults (Fung et al., 2018). Interestingly, in contrast to our findings, larger parahippocampal volume has been associated with increased somatic complaints, a core feature of anxiety, and somatic complaints were shown to be associated with neuroticism-anxiety, a subscale of the neuroticism factor of the NEO Personality Inventory, in individuals with increased parahippocampal gyrus volume (Wei et al., 2014).

Finally, the thalamus also showed a negative association with anxiety in Model 1 when SES was not included. Beyond its role in relaying sensory and motor signals and regulating consciousness and alertness (Torrico & Munakomi, 2025), the thalamus is implicated in chronic stress regulation and anxiety-like behaviours (Bhatnagar et al., 2000; 2003; Hsu et al., 2014; Li et al., 2010). Reduced thalamic GMV and density has been reported in patients with SAD and GAD (Meng et al., 2013; Moon et al., 2015; Wang et al., 2018; Zhao et al., 2017) along with abnormal asymmetry in thalamic volume in SAD (Zhang et al., 2020).

While these findings align with prior literature, most previous studies have focused on children or younger to middle-aged adults, and we have been being unable to find research highlighting a relationship between structure of the precentral gyrus, supramarginal gyrus, and thalamus, and minimal research for the insula (Potvin et al., 2015) and the parahippocampal gyrus (Pink et al., 2017) in anxiety in older adults, making comparisons difficult. However, given the lack of research of the relationship between anxiety and brain structure in this age group, this is unsurprising and further highlights the novelty and necessity of the current study in addressing gaps in our understanding of anxiety-related neuroanatomical differences in older adults.

### The effect of Education and Income

Several brain regions were significantly associated with anxiety in Model 1 when SES was not considered, but all but one of these relationships became smaller and non-significant when SES was included. There are several different and not mutually exclusive interpretations for why this might be. On the one hand there could be effects directly related to the predictors at the individual level, e.g. that the experience of spending longer in formal education has benefits for brain development and mental health. On the other hand, education and income could be acting as proxy variables for other, individual and/or social factors, such as predisposition to having higher educational achievement and therefore better paying employment, or differences in social environment associated with more education and income such as better housing, more green space, or less crime.

These findings vary in terms of their brain location and function, so interpretation of the results is more difficult, however they have all been related to SES in previous studies (Jednoróg et al., 2012; Jenkins et al., 2020; McDermott et al., 2019; Noble et al., 2015; Thanaraju et al., 2024). Also, given that SES affects whole brain health and development (Brito & Noble, 2014; Lu et al., 2021; Rakesh et al., 2023; Resende et al., 2019), it is unsurprising that varying regions were implicated.

The fact that SES affected the relationship between anxiety and supramarginal gyrus volume aligns with evidence linking education to right supramarginal gyrus volume and emotion regulation (Wada et al., 2021). Studies suggest that education contributes to preserving this region, indirectly influencing emotion recognition. Additionally, research on socioeconomic disadvantage and neural organisation found community-level SES affected left supramarginal gyrus connectivity, while household income showed no significant effect (Gellci et al., 2018). This supports the notion that family and neighbourhood poverty impact brain development differently, with education potentially playing a larger role than income (Qiu et al., 2025). Findings on SES and supramarginal gyrus structure are mixed. Noble et al. (2015) found parental education, but not income, was linked to surface area variation.

McDermott et al. (2019) reported a stable positive relationship between SES and right supramarginal gyrus cortical thickness in youth, while Romeo et al. (2018) found a similar association bilaterally in children with reading disabilities. Given this, SES-related preservation of supramarginal gyrus integrity may explain why the negative association between anxiety and volume was no longer significant when SES was included. The insula has also been associated with SES in existing literature. In addition to the findings in the supramarginal gyrus mentioned above, Romeo et al. (2018), found that higher SES, comprised of maternal education and occupational prestige was associated with increased cortical thickness of left insula in children aged 6-9 with reading disability. Noble et al. (2015) also found that parental education was significantly associated with variation in surface area in the insula bilaterally. In addition, Jednoróg et al. (2012) found significant positive correlations of GMV in the bilateral insula and SES as measured by maternal education and current profession when investigating the influence of SES on children’s brain structure, in a sample of 23 10-year-old children with a wide range of parental SES. Again, a positive relationship between SES and brain structure could explain why, when we considered SES, the negative relationship between anxiety and brain structure in this region was no longer significant; this relationship could have been confounded by the relationship between the volume of the insula and SES, whether that be education or income.

While there are fewer existing studies reporting an association between SES and structure of the parahippocampal gyrus, Jednoróg et al. (2012), in the same study reported above, found significant positive correlations of GMV in the bilateral parahippocampal gyri and SES as measured by maternal education and current profession. In addition, in a study examining the relationship between SES, comprised of parental education and household income, and parahippocampal cortical thickness and racial differences in these relationships, Darvishi et al. (2021) reported that high income was associated with increased bilateral parahippocampal cortical thickness, with no effect of ethnicity on this relationship. For parental education however, ethnicity significantly interacted with its association with parahippocampal cortical thickness. Specifically, that the relationship between higher parental education and increased parahippocampal cortical thickness was stronger for white rather than black and other/mixed-ethnicity pre-adolescents.

The thalamus was also shown to be affected by SES in the prior mentioned study by McDermott et al. (2019) that also found effects in the right supramarginal gyrus. They reported that higher childhood SES is associated with larger bilateral volumes of the thalamus and that this finding was stable between the ages of 5 and 25 and that this effect was the greatest of all subcortical structures. Loued-Khenissi et al. (2022) further reported that childhood SES is linked to right thalamic volume, while adult SES is associated with left thalamic volume.

Unlike the other regions, the precentral gyrus remained significantly associated with anxiety scores when controlling for SES. This suggests either that education and income do not strongly influence precentral gyrus volume or that their effect is insufficient to eliminate the observed anxiety-related associated in this region. Given prior evidence linking this region to SES (Dufford et al., 2021; Kim et al., 2022; Takeuchi et al., 2021; Tomasi & Volkow, 2021), the latter explanation seems more plausible.

Overall, these findings reinforce the interdependence of brain structure, anxiety and SES, highlighting the need to consider all three factors when examining these relationships. Neuroimaging studies examining anxiety and brain structure without accounting for social determinants may overlook critical influences in this relationship.

### Limitations

The current study was limited by the fact that it did not exclude participants with conditions that could affect the brain. However, given the large sample size and the fact that it is a general population study, it hopefully would not have affected the results systematically. We also did not control for depression, which is highly comorbid with anxiety, with 45.7% of respondents of a worldwide mental health survey with lifetime MDD also reporting one or more lifetime anxiety disorders (Kessler et al., 2015). In addition, around three quarters of participants in the UKB who met the criteria for lifetime anxiety also met the criteria for lifetime depression (Davis et al., 2020). Depression has also been shown to be associated with brain structure and socioeconomic status in the UKB (Harris et al., 2022; Johns et al., 2025; Lyall et al., 2025; Qi et al., 2024; Ye et al., 2021). However, the decision was taken to look at the construct of anxiety as a whole, and to not control for depression, as to control for it would be removing the part of anxiety that covaries with depression (and vice versa). The goal was to increase our understanding of the relationship between anxiety and brain structure individually and then lead the way for future research to examine models with and without depression, allowing for careful interpretation of the interplay of the two disorders, which is beyond the scope of the current study.

UKB is limited in its diversity in terms of ethnicity, age and income and has a healthy-volunteer selection bias which leads it to not be representative on a number of different sociodemographic, physical, lifestyle and health-related variables (Fry et al., 2017), which could suggest that the sample may not be particularly anxious. In terms of anxiety however, Davis et al. (2020) compared rates in the UKB to those in the Health Survey England (HSE), a yearly face-to-face household survey and found that the rates in the UKB were higher than those in the 2014 HSE, which included 8000 adult participants designed to be representative of the adult population of the UK. 14% of participants in the UKB were categorised as having anxiety, nerves or GAD with an additional 1.2% having social anxiety or social phobia. In the HSE, 5.2% were categorised as having GAD, they were not asked about general anxiety or worry, which Davis et al. (2020) suggested might explain the differences in rates. In addition, the present study used anxiety scores on a continuous scale and did not factor in whether individuals had a diagnosis or met the diagnostic cut-off for anxiety disorders. However, there is existing evidence of a relationship between subthreshold anxiety and brain structure (Besteher et al., 2020; Kim et al., 2023; Machado-de-Sousa et al., 2014) and as significant relationships were found in this study, this does not detract from the results found here.

Another potential limitation is that the time between the online questionnaires including the questionnaires about anxiety and sociodemographic information and the brain imaging scans varied across participants. Dutt et al. (2020) reported timing effects between scan acquisition and online questionnaire which they stated was highly inconsistent across participants, which may be a source of error in our study when comparing current anxiety symptoms to brain structure.

Finally, as this study was cross-sectional and the UKB does not include information on an individual’s past SES, we do not have information on the participants’ socioeconomic circumstances when their brains were developing. For example, individuals could have high SES now but have experienced deprivation during the “critical period” of development. There is evidence that experience socioeconomic disadvantage during childhood or adolescence, which are key periods of brain development (Cisneros-Franco et al., 2020; Gale, 2004; Konrad et al., 2013; Larsen & Luna, 2018), can have a negative impact on brain health in later life, irrespective of future SES (Thanaraju et al., 2024). However, there is some evidence that higher SES may be able to alleviate some of the damage caused by lower SES in childhood (Tharanju et al., 2024). It could be argued that education may be less at risk of the effects of this, as past a certain point it does not change, however income could be at risk of being affected by this.

### Future Work

Given the limitations highlighted, there is a need for large-scale longitudinal research to clarify the causal relationships between the factors examined in this study and to gain a deeper understanding of the underlying mechanisms. A crucial next step is to examine how socioeconomic factors combine with structural brain correlates to predict anxiety, allowing for a clearer delineation of the relative contribution of socioeconomic status and brain structure to anxiety. Longitudinal studies tracking SES alongside changes in anxiety symptoms and brain structure over time would provide valuable insight into how these factors influence one another and how their relationships evolve as anxiety symptoms fluctuate. It remains unclear whether education and income have a direct causal impact on brain structure or if they serve as proxy measures for other factors that shape brain development and structural changes across the lifespan. A comprehensive understanding of these relationships is essential for generating evidence that can inform policies aimed at reducing health inequalities and mitigating the well-documented effects of anxiety on individuals and society

### Conclusion

To our knowledge, this is the first study to investigate the relationship between anxiety, cortical and subcortical volume, and socioeconomic status (SES) in a middle-aged to older adult sample of this size and adds to the limited literature examining the relationship between anxiety and brain structure in this age group. Given the influence of SES on the association between anxiety and brain structure, we propose that SES should be an essential consideration for clinicians and researchers working with neuroimaging data in this area. Future large-scale longitudinal research is essential to establish the causal relationships among the factors explored in this study.

## Data Availability

Data in the present study are available from the UK Bioabank to approved researchers

## Supplementary Materials

**Table S1.**
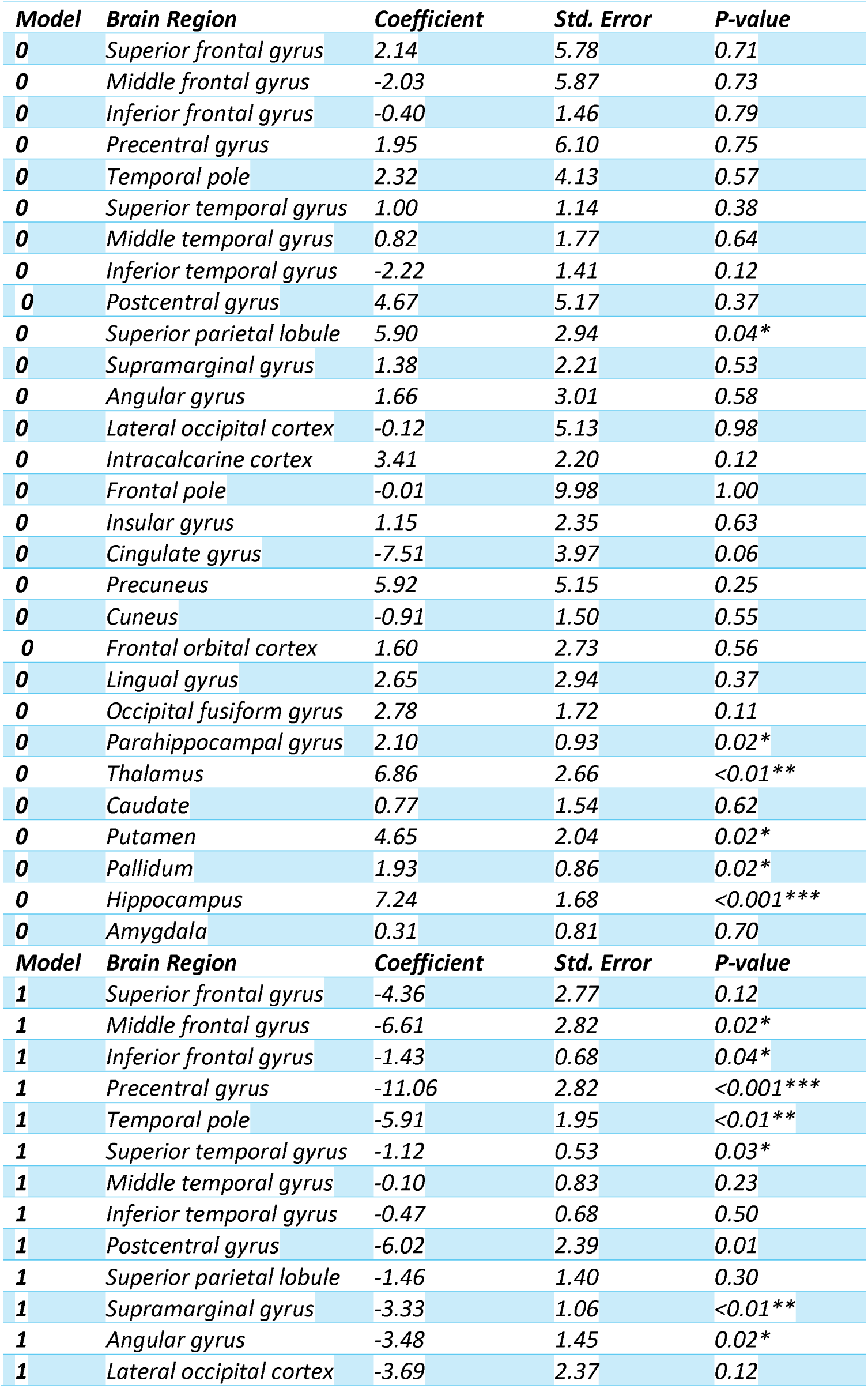
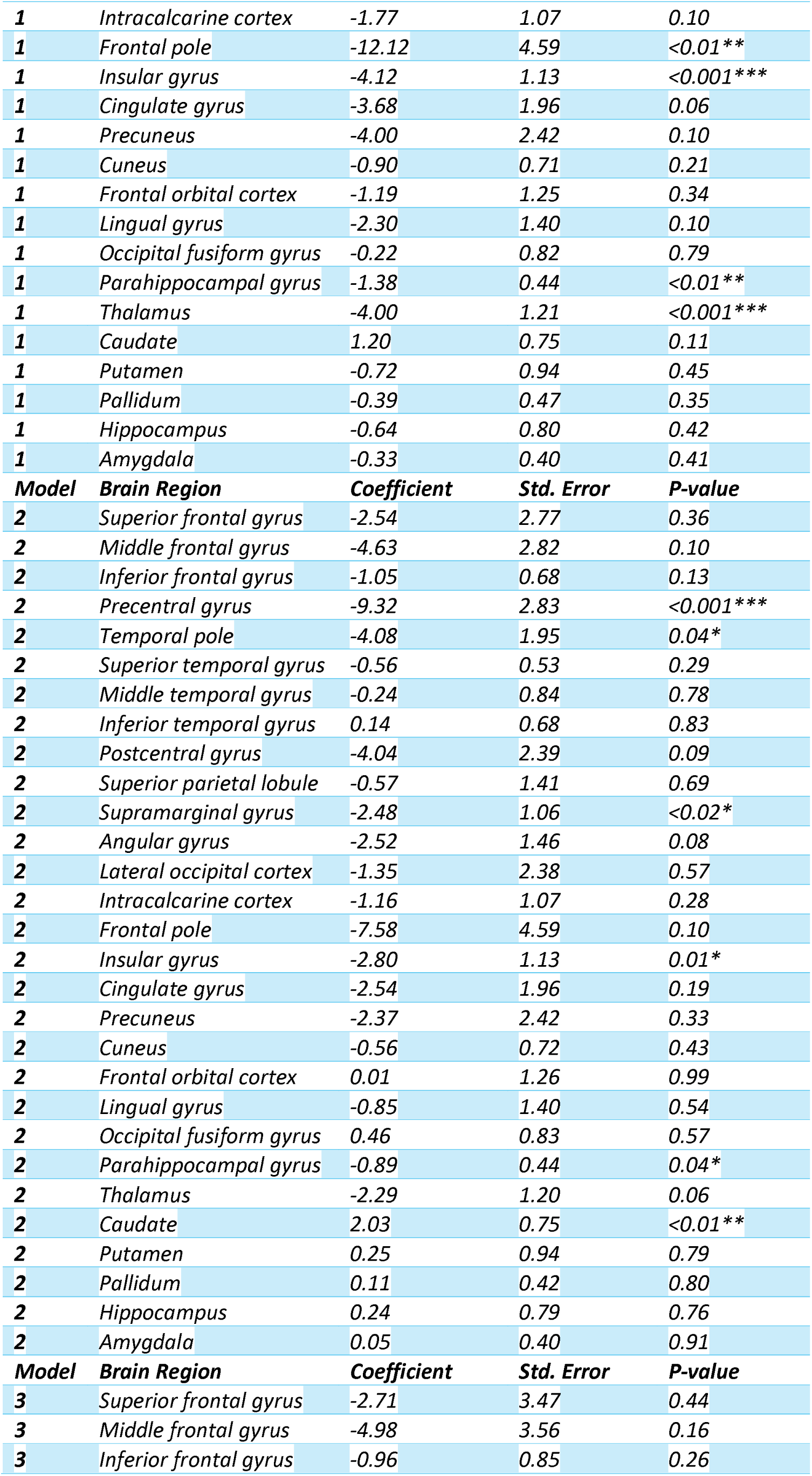
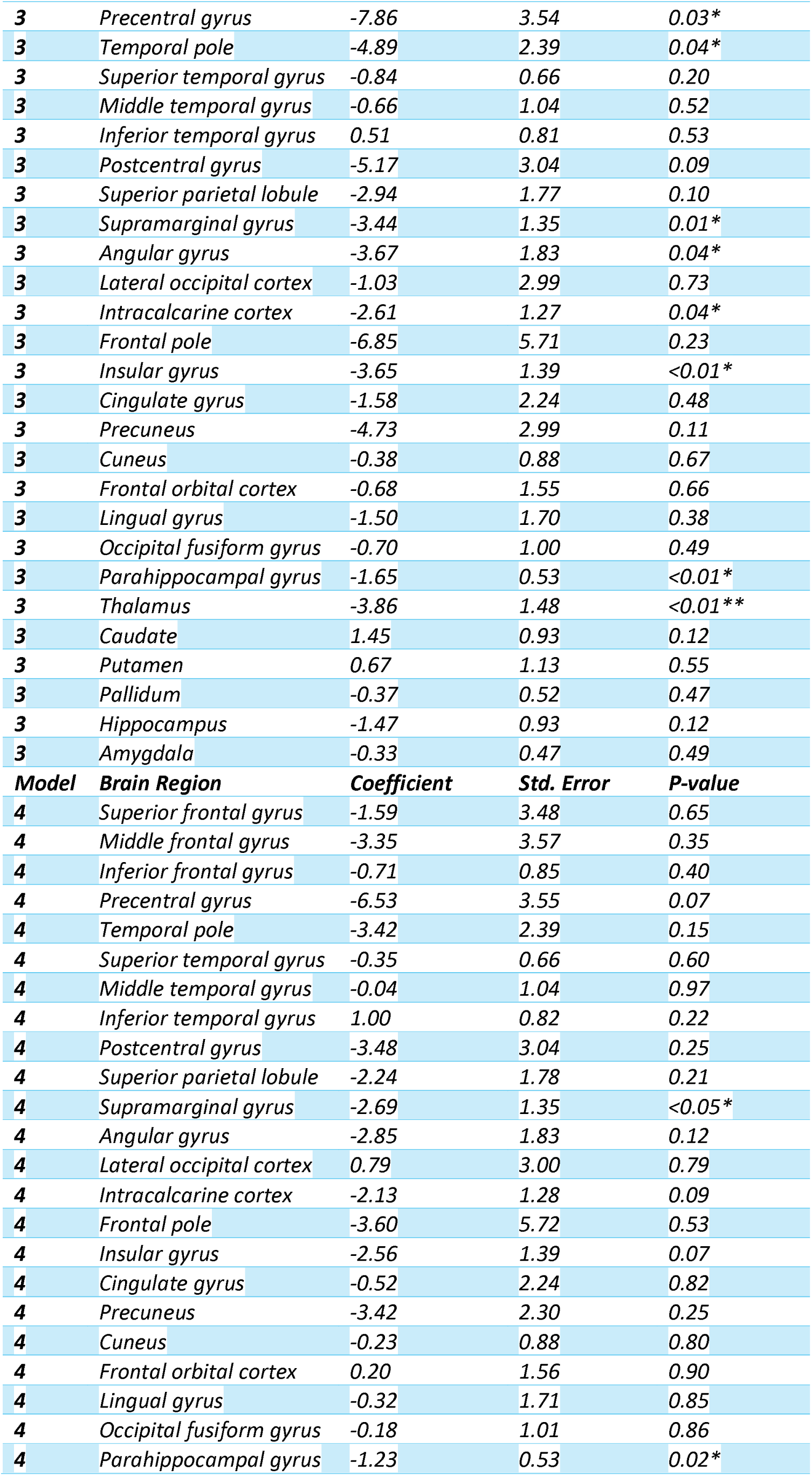
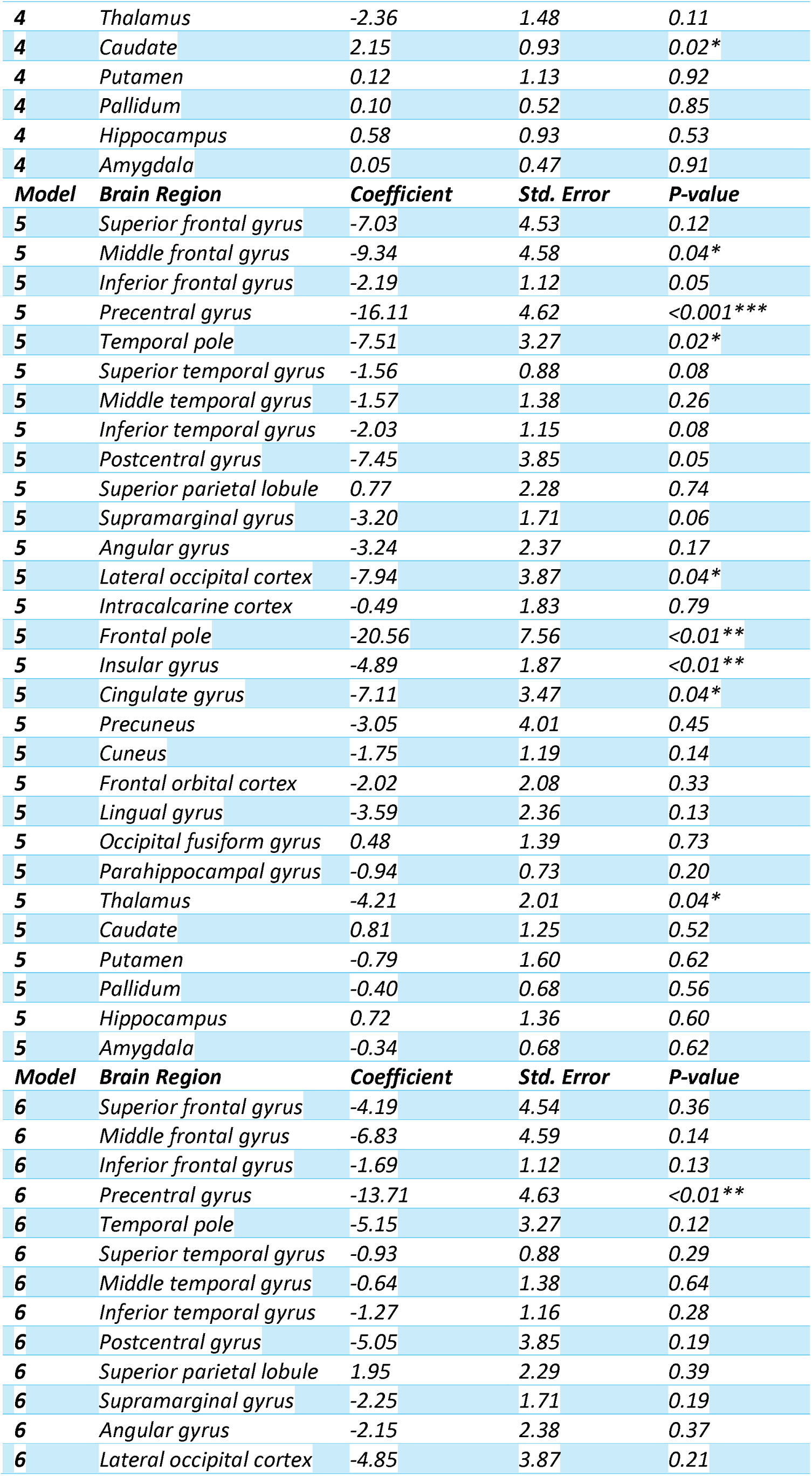
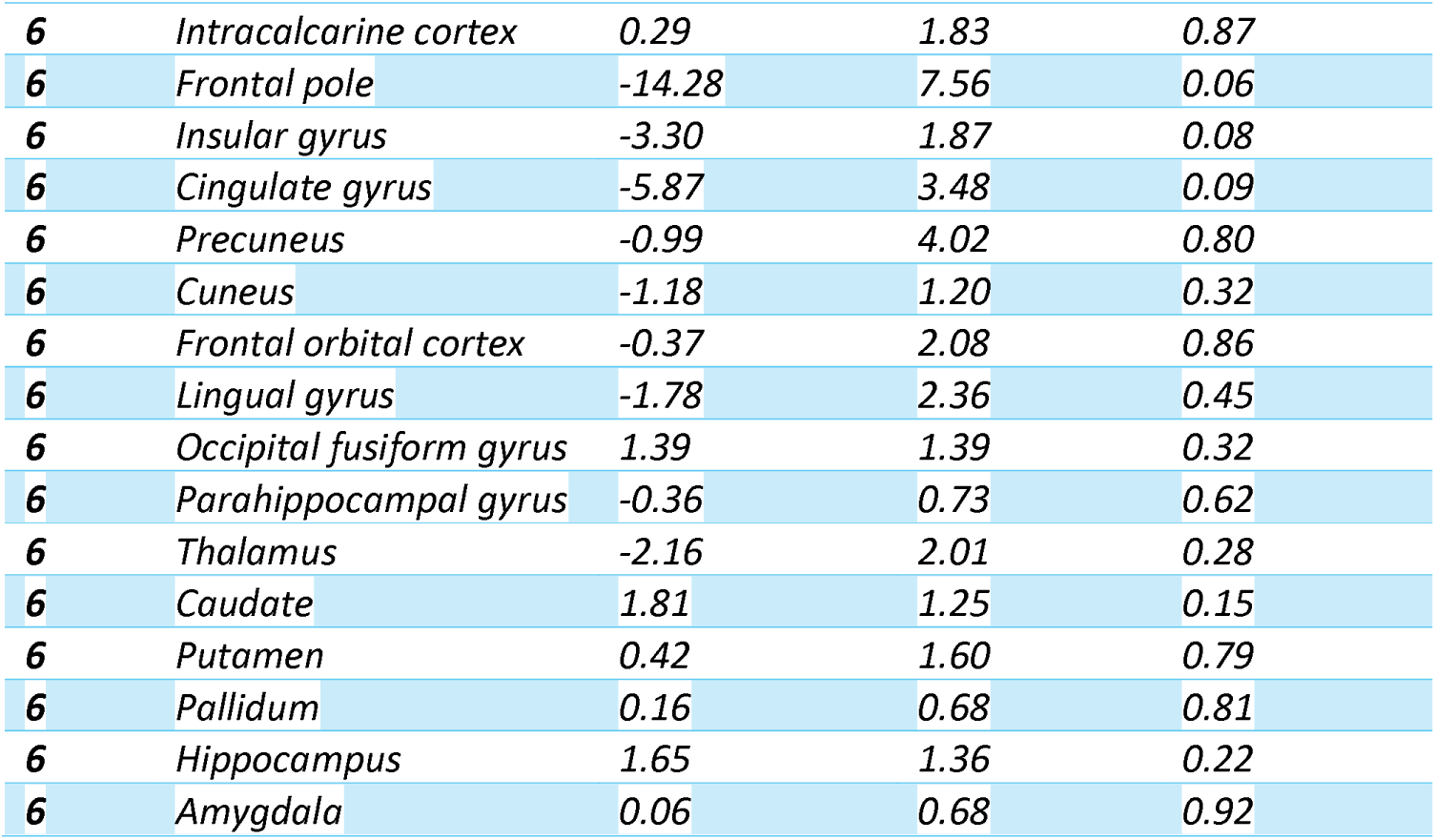
Results for each of the multivariate regression analysis models.

