## Supplemental Table 1 for "Anxiety, brain structure and socioeconomic status in middle-aged and older adults"

**Supplementary Materials**

*Table S1 Results for each of the multivariate regression analysis models*

| *Model* | *Brain Region* | *Coefficient* | *Std. Error* | *P-value* |
| --- | --- | --- | --- | --- |
| *0* | *Superior frontal gyrus* | *2.14* | *5.78* | *0.71* |
| *0* | *Middle frontal gyrus* | *-2.03* | *5.87* | *0.73* |
| *0* | *Inferior frontal gyrus* | *-0.40* | *1.46* | *0.79* |
| *0* | *Precentral gyrus* | *1.95* | *6.10* | *0.75* |
| *0* | *Temporal pole* | *2.32* | *4.13* | *0.57* |
| *0* | *Superior temporal gyrus* | *1.00* | *1.14* | *0.38* |
| *0* | *Middle temporal gyrus* | *0.82* | *1.77* | *0.64* |
| *0* | *Inferior temporal gyrus* | *-2.22* | *1.41* | *0.12* |
| *0* | *Postcentral gyrus* | *4.67* | *5.17* | *0.37* |
| *0* | *Superior parietal lobule* | *5.90* | *2.94* | *0.04** |
| *0* | *Supramarginal gyrus* | *1.38* | *2.21* | *0.53* |
| *0* | *Angular gyrus* | *1.66* | *3.01* | *0.58* |
| *0* | *Lateral occipital cortex* | *-0.12* | *5.13* | *0.98* |
| *0* | *Intracalcarine cortex* | *3.41* | *2.20* | *0.12* |
| *0* | *Frontal pole* | *-0.01* | *9.98* | *1.00* |
| *0* | *Insular gyrus* | *1.15* | *2.35* | *0.63* |
| *0* | *Cingulate gyrus* | *-7.51* | *3.97* | *0.06* |
| *0* | *Precuneus* | *5.92* | *5.15* | *0.25* |
| *0* | *Cuneus* | *-0.91* | *1.50* | *0.55* |
| *0* | *Frontal orbital cortex* | *1.60* | *2.73* | *0.56* |
| *0* | *Lingual gyrus* | *2.65* | *2.94* | *0.37* |
| *0* | *Occipital fusiform gyrus* | *2.78* | *1.72* | *0.11* |
| *0* | *Parahippocampal gyrus* | *2.10* | *0.93* | *0.02** |
| *0* | *Thalamus* | *6.86* | *2.66* | *<0.01*** |
| *0* | *Caudate* | *0.77* | *1.54* | *0.62* |
| *0* | *Putamen* | *4.65* | *2.04* | *0.02** |
| *0* | *Pallidum* | *1.93* | *0.86* | *0.02** |
| *0* | *Hippocampus* | *7.24* | *1.68* | *<0.001**** |
| *0* | *Amygdala* | *0.31* | *0.81* | *0.70* |
| *Model* | ***Brain Region*** | ***Coefficient*** | ***Std. Error*** | ***P-value*** |
| *1* | *Superior frontal gyrus* | *-4.36* | *2.77* | *0.12* |
| *1* | *Middle frontal gyrus* | *-6.61* | *2.82* | *0.02** |
| *1* | *Inferior frontal gyrus* | *-1.43* | *0.68* | *0.04** |
| *1* | *Precentral gyrus* | *-11.06* | *2.82* | *<0.001**** |
| *1* | *Temporal pole* | *-5.91* | *1.95* | *<0.01*** |
| *1* | *Superior temporal gyrus* | *-1.12* | *0.53* | *0.03** |
| *1* | *Middle temporal gyrus* | *-0.10* | *0.83* | *0.23* |
| *1* | *Inferior temporal gyrus* | *-0.47* | *0.68* | *0.50* |
| *1* | *Postcentral gyrus* | *-6.02* | *2.39* | *0.01* |
| *1* | *Superior parietal lobule* | *-1.46* | *1.40* | *0.30* |
| *1* | *Supramarginal gyrus* | *-3.33* | *1.06* | *<0.01*** |
| *1* | *Angular gyrus* | *-3.48* | *1.45* | *0.02** |
| *1* | *Lateral occipital cortex* | *-3.69* | *2.37* | *0.12* |
| *1* | *Intracalcarine cortex* | *-1.77* | *1.07* | *0.10* |
| *1* | *Frontal pole* | *-12.12* | *4.59* | *<0.01*** |
| *1* | *Insular gyrus* | *-4.12* | *1.13* | *<0.001**** |
| *1* | *Cingulate gyrus* | *-3.68* | *1.96* | *0.06* |
| *1* | *Precuneus* | *-4.00* | *2.42* | *0.10* |
| *1* | *Cuneus* | *-0.90* | *0.71* | *0.21* |
| *1* | *Frontal orbital cortex* | *-1.19* | *1.25* | *0.34* |
| *1* | *Lingual gyrus* | *-2.30* | *1.40* | *0.10* |
| *1* | *Occipital fusiform gyrus* | *-0.22* | *0.82* | *0.79* |
| *1* | *Parahippocampal gyrus* | *-1.38* | *0.44* | *<0.01*** |
| *1* | *Thalamus* | *-4.00* | *1.21* | *<0.001**** |
| *1* | *Caudate* | *1.20* | *0.75* | *0.11* |
| *1* | *Putamen* | *-0.72* | *0.94* | *0.45* |
| *1* | *Pallidum* | *-0.39* | *0.47* | *0.35* |
| *1* | *Hippocampus* | *-0.64* | *0.80* | *0.42* |
| *1* | *Amygdala* | *-0.33* | *0.40* | *0.41* |
| *Model* | ***Brain Region*** | ***Coefficient*** | ***Std. Error*** | ***P-value*** |
| *2* | *Superior frontal gyrus* | *-2.54* | *2.77* | *0.36* |
| *2* | *Middle frontal gyrus* | *-4.63* | *2.82* | *0.10* |
| *2* | *Inferior frontal gyrus* | *-1.05* | *0.68* | *0.13* |
| *2* | *Precentral gyrus* | *-9.32* | *2.83* | *<0.001**** |
| *2* | *Temporal pole* | *-4.08* | *1.95* | *0.04** |
| *2* | *Superior temporal gyrus* | *-0.56* | *0.53* | *0.29* |
| *2* | *Middle temporal gyrus* | *-0.24* | *0.84* | *0.78* |
| *2* | *Inferior temporal gyrus* | *0.14* | *0.68* | *0.83* |
| *2* | *Postcentral gyrus* | *-4.04* | *2.39* | *0.09* |
| *2* | *Superior parietal lobule* | *-0.57* | *1.41* | *0.69* |
| *2* | *Supramarginal gyrus* | *-2.48* | *1.06* | *<0.02** |
| *2* | *Angular gyrus* | *-2.52* | *1.46* | *0.08* |
| *2* | *Lateral occipital cortex* | *-1.35* | *2.38* | *0.57* |
| *2* | *Intracalcarine cortex* | *-1.16* | *1.07* | *0.28* |
| *2* | *Frontal pole* | *-7.58* | *4.59* | *0.10* |
| *2* | *Insular gyrus* | *-2.80* | *1.13* | *0.01** |
| *2* | *Cingulate gyrus* | *-2.54* | *1.96* | *0.19* |
| *2* | *Precuneus* | *-2.37* | *2.42* | *0.33* |
| *2* | *Cuneus* | *-0.56* | *0.72* | *0.43* |
| *2* | *Frontal orbital cortex* | *0.01* | *1.26* | *0.99* |
| *2* | *Lingual gyrus* | *-0.85* | *1.40* | *0.54* |
| *2* | *Occipital fusiform gyrus* | *0.46* | *0.83* | *0.57* |
| *2* | *Parahippocampal gyrus* | *-0.89* | *0.44* | *0.04** |
| *2* | *Thalamus* | *-2.29* | *1.20* | *0.06* |
| *2* | *Caudate* | *2.03* | *0.75* | *<0.01*** |
| *2* | *Putamen* | *0.25* | *0.94* | *0.79* |
| *2* | *Pallidum* | *0.11* | *0.42* | *0.80* |
| *2* | *Hippocampus* | *0.24* | *0.79* | *0.76* |
| *2* | *Amygdala* | *0.05* | *0.40* | *0.91* |
| *Model* | ***Brain Region*** | ***Coefficient*** | ***Std. Error*** | ***P-value*** |
| *3* | *Superior frontal gyrus* | *-2.71* | *3.47* | *0.44* |
| *3* | *Middle frontal gyrus* | *-4.98* | *3.56* | *0.16* |
| *3* | *Inferior frontal gyrus* | *-0.96* | *0.85* | *0.26* |
| *3* | *Precentral gyrus* | *-7.86* | *3.54* | *0.03** |
| *3* | *Temporal pole* | *-4.89* | *2.39* | *0.04** |
| *3* | *Superior temporal gyrus* | *-0.84* | *0.66* | *0.20* |
| *3* | *Middle temporal gyrus* | *-0.66* | *1.04* | *0.52* |
| *3* | *Inferior temporal gyrus* | *0.51* | *0.81* | *0.53* |
| *3* | *Postcentral gyrus* | *-5.17* | *3.04* | *0.09* |
| *3* | *Superior parietal lobule* | *-2.94* | *1.77* | *0.10* |
| *3* | *Supramarginal gyrus* | *-3.44* | *1.35* | *0.01** |
| *3* | *Angular gyrus* | *-3.67* | *1.83* | *0.04** |
| *3* | *Lateral occipital cortex* | *-1.03* | *2.99* | *0.73* |
| *3* | *Intracalcarine cortex* | *-2.61* | *1.27* | *0.04** |
| *3* | *Frontal pole* | *-6.85* | *5.71* | *0.23* |
| *3* | *Insular gyrus* | *-3.65* | *1.39* | *<0.01** |
| *3* | *Cingulate gyrus* | *-1.58* | *2.24* | *0.48* |
| *3* | *Precuneus* | *-4.73* | *2.99* | *0.11* |
| *3* | *Cuneus* | *-0.38* | *0.88* | *0.67* |
| *3* | *Frontal orbital cortex* | *-0.68* | *1.55* | *0.66* |
| *3* | *Lingual gyrus* | *-1.50* | *1.70* | *0.38* |
| *3* | *Occipital fusiform gyrus* | *-0.70* | *1.00* | *0.49* |
| *3* | *Parahippocampal gyrus* | *-1.65* | *0.53* | *<0.01** |
| *3* | *Thalamus* | *-3.86* | *1.48* | *<0.01*** |
| *3* | *Caudate* | *1.45* | *0.93* | *0.12* |
| *3* | *Putamen* | *0.67* | *1.13* | *0.55* |
| *3* | *Pallidum* | *-0.37* | *0.52* | *0.47* |
| *3* | *Hippocampus* | *-1.47* | *0.93* | *0.12* |
| *3* | *Amygdala* | *-0.33* | *0.47* | *0.49* |
| *Model* | ***Brain Region*** | ***Coefficient*** | ***Std. Error*** | ***P-value*** |
| *4* | *Superior frontal gyrus* | *-1.59* | *3.48* | *0.65* |
| *4* | *Middle frontal gyrus* | *-3.35* | *3.57* | *0.35* |
| *4* | *Inferior frontal gyrus* | *-0.71* | *0.85* | *0.40* |
| *4* | *Precentral gyrus* | *-6.53* | *3.55* | *0.07* |
| *4* | *Temporal pole* | *-3.42* | *2.39* | *0.15* |
| *4* | *Superior temporal gyrus* | *-0.35* | *0.66* | *0.60* |
| *4* | *Middle temporal gyrus* | *-0.04* | *1.04* | *0.97* |
| *4* | *Inferior temporal gyrus* | *1.00* | *0.82* | *0.22* |
| *4* | *Postcentral gyrus* | *-3.48* | *3.04* | *0.25* |
| *4* | *Superior parietal lobule* | *-2.24* | *1.78* | *0.21* |
| *4* | *Supramarginal gyrus* | *-2.69* | *1.35* | *<0.05** |
| *4* | *Angular gyrus* | *-2.85* | *1.83* | *0.12* |
| *4* | *Lateral occipital cortex* | *0.79* | *3.00* | *0.79* |
| *4* | *Intracalcarine cortex* | *-2.13* | *1.28* | *0.09* |
| *4* | *Frontal pole* | *-3.60* | *5.72* | *0.53* |
| *4* | *Insular gyrus* | *-2.56* | *1.39* | *0.07* |
| *4* | *Cingulate gyrus* | *-0.52* | *2.24* | *0.82* |
| *4* | *Precuneus* | *-3.42* | *2.30* | *0.25* |
| *4* | *Cuneus* | *-0.23* | *0.88* | *0.80* |
| *4* | *Frontal orbital cortex* | *0.20* | *1.56* | *0.90* |
| *4* | *Lingual gyrus* | *-0.32* | *1.71* | *0.85* |
| *4* | *Occipital fusiform gyrus* | *-0.18* | *1.01* | *0.86* |
| *4* | *Parahippocampal gyrus* | *-1.23* | *0.53* | *0.02** |
| *4* | *Thalamus* | *-2.36* | *1.48* | *0.11* |
| *4* | *Caudate* | *2.15* | *0.93* | *0.02** |
| *4* | *Putamen* | *0.12* | *1.13* | *0.92* |
| *4* | *Pallidum* | *0.10* | *0.52* | *0.85* |
| *4* | *Hippocampus* | *0.58* | *0.93* | *0.53* |
| *4* | *Amygdala* | *0.05* | *0.47* | *0.91* |
| *Model* | ***Brain Region*** | ***Coefficient*** | ***Std. Error*** | ***P-value*** |
| *5* | *Superior frontal gyrus* | *-7.03* | *4.53* | *0.12* |
| *5* | *Middle frontal gyrus* | *-9.34* | *4.58* | *0.04** |
| *5* | *Inferior frontal gyrus* | *-2.19* | *1.12* | *0.05* |
| *5* | *Precentral gyrus* | *-16.11* | *4.62* | *<0.001**** |
| *5* | *Temporal pole* | *-7.51* | *3.27* | *0.02** |
| *5* | *Superior temporal gyrus* | *-1.56* | *0.88* | *0.08* |
| *5* | *Middle temporal gyrus* | *-1.57* | *1.38* | *0.26* |
| *5* | *Inferior temporal gyrus* | *-2.03* | *1.15* | *0.08* |
| *5* | *Postcentral gyrus* | *-7.45* | *3.85* | *0.05* |
| *5* | *Superior parietal lobule* | *0.77* | *2.28* | *0.74* |
| *5* | *Supramarginal gyrus* | *-3.20* | *1.71* | *0.06* |
| *5* | *Angular gyrus* | *-3.24* | *2.37* | *0.17* |
| *5* | *Lateral occipital cortex* | *-7.94* | *3.87* | *0.04** |
| *5* | *Intracalcarine cortex* | *-0.49* | *1.83* | *0.79* |
| *5* | *Frontal pole* | *-20.56* | *7.56* | *<0.01*** |
| *5* | *Insular gyrus* | *-4.89* | *1.87* | *<0.01*** |
| *5* | *Cingulate gyrus* | *-7.11* | *3.47* | *0.04** |
| *5* | *Precuneus* | *-3.05* | *4.01* | *0.45* |
| *5* | *Cuneus* | *-1.75* | *1.19* | *0.14* |
| *5* | *Frontal orbital cortex* | *-2.02* | *2.08* | *0.33* |
| *5* | *Lingual gyrus* | *-3.59* | *2.36* | *0.13* |
| *5* | *Occipital fusiform gyrus* | *0.48* | *1.39* | *0.73* |
| *5* | *Parahippocampal gyrus* | *-0.94* | *0.73* | *0.20* |
| *5* | *Thalamus* | *-4.21* | *2.01* | *0.04** |
| *5* | *Caudate* | *0.81* | *1.25* | *0.52* |
| *5* | *Putamen* | *-0.79* | *1.60* | *0.62* |
| *5* | *Pallidum* | *-0.40* | *0.68* | *0.56* |
| *5* | *Hippocampus* | *0.72* | *1.36* | *0.60* |
| *5* | *Amygdala* | *-0.34* | *0.68* | *0.62* |
| *Model* | ***Brain Region*** | ***Coefficient*** | ***Std. Error*** | ***P-value*** |
| *6* | *Superior frontal gyrus* | *-4.19* | *4.54* | *0.36* |
| *6* | *Middle frontal gyrus* | *-6.83* | *4.59* | *0.14* |
| *6* | *Inferior frontal gyrus* | *-1.69* | *1.12* | *0.13* |
| *6* | *Precentral gyrus* | *-13.71* | *4.63* | *<0.01*** |
| *6* | *Temporal pole* | *-5.15* | *3.27* | *0.12* |
| *6* | *Superior temporal gyrus* | *-0.93* | *0.88* | *0.29* |
| *6* | *Middle temporal gyrus* | *-0.64* | *1.38* | *0.64* |
| *6* | *Inferior temporal gyrus* | *-1.27* | *1.16* | *0.28* |
| *6* | *Postcentral gyrus* | *-5.05* | *3.85* | *0.19* |
| *6* | *Superior parietal lobule* | *1.95* | *2.29* | *0.39* |
| *6* | *Supramarginal gyrus* | *-2.25* | *1.71* | *0.19* |
| *6* | *Angular gyrus* | *-2.15* | *2.38* | *0.37* |
| *6* | *Lateral occipital cortex* | *-4.85* | *3.87* | *0.21* |
| *6* | *Intracalcarine cortex* | *0.29* | *1.83* | *0.87* |
| *6* | *Frontal pole* | *-14.28* | *7.56* | *0.06* |
| *6* | *Insular gyrus* | *-3.30* | *1.87* | *0.08* |
| *6* | *Cingulate gyrus* | *-5.87* | *3.48* | *0.09* |
| *6* | *Precuneus* | *-0.99* | *4.02* | *0.80* |
| *6* | *Cuneus* | *-1.18* | *1.20* | *0.32* |
| *6* | *Frontal orbital cortex* | *-0.37* | *2.08* | *0.86* |
| *6* | *Lingual gyrus* | *-1.78* | *2.36* | *0.45* |
| *6* | *Occipital fusiform gyrus* | *1.39* | *1.39* | *0.32* |
| *6* | *Parahippocampal gyrus* | *-0.36* | *0.73* | *0.62* |
| *6* | *Thalamus* | *-2.16* | *2.01* | *0.28* |
| *6* | *Caudate* | *1.81* | *1.25* | *0.15* |
| *6* | *Putamen* | *0.42* | *1.60* | *0.79* |
| *6* | *Pallidum* | *0.16* | *0.68* | *0.81* |
| *6* | *Hippocampus* | *1.65* | *1.36* | *0.22* |
| *6* | *Amygdala* | *0.06* | *0.68* | *0.92* |
